# Is Catheter Ablation Associated with Preservation of Cognitive Function? An Analysis From the SAGE-AF Observational Cohort Study

**DOI:** 10.1101/2023.11.20.23298768

**Authors:** Bahadar S. Srichawla, Alexander P. Hamel, Philip Cook, Rozaleen Aleyadeh, Darleen Lessard, Edith M. Otabil, Jordy Mehawej, Jane S. Saczynski, David D. McManus, Majaz Moonis

## Abstract

**Objectives:** To examine the associations between catheter ablation treatment (CA) versus medical management and cognitive impairment among older adults with atrial fibrillation (AF).

**Methods:** Ambulatory patients who had AF, were ≥ 65-years-old, and were eligible to receive oral anticoagulation could be enrolled into the SAGE (Systematic Assessment of Geriatric Elements)-AF study from internal medicine and cardiology clinics in Massachusetts and Georgia between 2016 and 2018. Cognitive function was assessed using the Montreal Cognitive Assessment (MoCA) tool at baseline, one-, and two years. Cognitive impairment was defined as a MoCA score ≤ 23. Multivariate-adjusted logistic regression of longitudinal repeated measures was used to examine associations between treatment with CA vs. medical management and cognitive impairment.

**Results:** 887 participants were included in this analysis. On average, participants were 75.2 ± 6.7 years old, 48.6% women, and 87.4% white non-Hispanic. 193 (21.8%) participants received a CA before enrollment. Participants who had previously undergone CA were significantly less likely to be cognitively impaired during the two-year study period (*aOR 0.70, 95% CI 0.50-0.97*) than those medically managed (i.e., rate and/or rhythm control), even after adjusting with propensity score for CA. At the two-year follow-up a significantly greater number of individuals in the non-CA group were cognitively impaired (MoCA ≤ 23) compared to the CA-group (311 [44.8%] vs. 58 [30.1%], *p=0.0002*).

**Conclusions:** In this two-year longitudinal prospective cohort study participants who underwent CA for AF before enrollment were less likely to have cognitive impairment than those who had not undergone CA.

## INTRODUCTION

Atrial fibrillation (AF) is the most common cardiac arrhythmia in the United States, affecting 3 to 6 million people [1]. The Framingham Heart Study demonstrated a 3-fold increase in the prevalence of AF in the past 50 years [2]. AF carries a two-fold increased risk of mortality and a five-fold risk of ischemic stroke. AF has also been associated with ischemic phenomena and cardioembolic-mediated infarction to critical organs including the gastrointestinal tract and kidneys, among others [3]. To mitigate the risk of stroke secondary to AF, anticoagulants (ACs) such as warfarin and direct oral anticoagulant (DOAC) therapy are often used based on the CHA_2_DS_2_-VASc score. The CHA_2_DS_2_-VASc score is a clinical prediction rule for estimating the risk of stroke in patients with non-rheumatic atrial fibrillation (AF), a common and irregular heart rhythm. However, the utilization of AC is not without risk. Major hemorrhagic events, including intracerebral hemorrhage (ICH), and gastrointestinal bleeding, are of concern [4].

Previous studies have shown that AF is independently associated with cognitive decline. Proposed pathophysiologic mechanisms include silent ischemic or hemorrhagic cerebral micro-infarction and/or impaired cerebral blood flow [5]. Catheter ablation (CA) has become a common rhythm control management strategy for patients with AF and is the most frequently performed cardiac ablation procedure worldwide. Restoring sinus rhythm via CA addresses several of the proposed pathophysiologic mechanisms that may link AF to cognitive decline. Bodagh et al. performed a systematic review that identified 10 studies that assessed cognitive function in patients who underwent CA versus medical management. Incident dementia was lower among patients who received CA for rhythm control of AF as compared to those treated with medicine. However, other studies involving patients with AF with a more rigorous longitudinal assessment of cognitive function have shown no relation between CA and cognitive decline [6]. Finally, one of the major confounders when examining the relations between CA and AF is the use of anticoagulation, which can also have important effects on cerebrovascular health. In addition to the clear reduction in ischemic stroke attributable to anticoagulation, its use has been shown to cause cerebral microbleeds (CMBs) which may reduce cognitive function [7, 8].

In this retrospective analysis, we used data from the Systematic Assessment of Geriatric Elements in Atrial Fibrillation (or SAGE-AF) study, a prospective cohort study that enrolled individuals with AF aged ≥ 65 years and followed them over two years to understand the relations between oral anticoagulation, cognitive and physical function. To better understand the potential links between CA and cognitive health, we compared cognitive function at baseline and 1- and 2-year follow-up examinations among those SAGE-AF participants who underwent CA with those treated medically (either rate or rhythm control).

## METHODS

### 2.1 Design, Setting, and Participants

We examined data from the prospective cohort multicenter SAGE-AF study, whose details have been previously described [9]. Briefly, the SAGE-AF study enrolled patients age ≥ 65 from ambulatory care sites in Massachusetts and Georgia from 2016 to 2018. Patients were eligible if they had a documented diagnosis of AF and a CHA_2_DS_2_-VASc (congestive heart failure, hypertension, age ≥75, diabetes, stroke, vascular disease, age 65-74, and sex) score ≥ 2. Exclusion criteria included being contraindicated for oral anticoagulants, being unable to provide signed informed consent in English, and being unable to participate in follow-up visits during the 2-year study period. Contraindications to anticoagulants included recent or planned major surgery, recent or active major bleeding, use of anticoagulants for reasons other than AF, or significant thrombocytopenia (platelet count <50,000).

All participants enrolled in the study provided their informed written consent for protocols approved by the institutional review boards of the University of Massachusetts Medical School, Boston University, and Mercer University. Each participant underwent a routine physical examination, which included an electrocardiogram (ECG) at baseline and 2 years. Participants also had their medical history abstracted in addition to a 60-minute computer-assisted interview that assessed mood, cognition, social support, and other key patient-reported measures using standardized measures. We specifically extracted age, sex, race, education level, cognitive ability, frailty, depression, anxiety, social isolation, and smoking status at baseline. Other factors assessed included a history of intracranial hemorrhage, GI bleeding, major bleeding, heart failure, coronary artery disease – myocardial infarction or angina, peripheral vascular disease, hypertension, type II diabetes, dyslipidemia, ischemic stroke, anemia, chronic obstructive pulmonary disease, renal disease, implantable cardiac device, and sleep apnea. Laboratory values extracted included creatinine, hemoglobin, and platelet count. The HAS-BLED score is a clinical prediction rule used to estimate the risk of major bleeding for patients who are on anticoagulation therapy, especially those with atrial fibrillation (AF). The CHA_2_DS_2_-VASc and HAS-BLED scores were calculated, and polypharmacy, utilization of anticoagulants, antiplatelet agents, and antiarrhythmics were assessed.

### 2.2 Global cognitive function assessment

Our primary outcome, cognitive function, was measured using the validated Montreal Cognitive Assessment (MoCA) tool, a 10-minute and 30-item screening tool used to detect cognitive impairment [10]. MoCA tests memory, visuospatial ability, executive function, attention, concentration, working memory, and orientation and is accepted for use in an older population of patients with strokes [11]. Cognitive impairment was defined as a cut-off of ≤ 23, in accordance with prior studies [12, 13]. Each participant included in this analysis completed the MoCA at baseline, 1-year, and 2-years.

### 2.3 AF Treatment

As part of the medical record abstraction, study coordinators collected procedural histories of participants, including prior ablation and medication use. Participants who had previously undergone a catheter ablation prior to their baseline SAGE-AF examination were considered to have undergone CA. 29 participants were ablated before their baseline exam and received another CA during the follow-up period. The specific type of CA performed was not reported, but since all CA participants were enrolled between 2016-2018 from US-based clinics, the majority of CA performed during this time were likely to have been radiofrequency ablation pulmonary vein isolation procedures [14].

For each participant, medications were abstracted from the electronic medical record and confirmed with the participant during the in-person interview. Relevant medications abstracted from the health record included OAC as well as rate and rhythm control agents. Specifically, prescriptions of antiarrhythmic, rate control, antiplatelet, and anticoagulant agents (including vitamin K antagonists and DOACs) were abstracted and confirmed with participants. Participants who did not receive CA were considered as ‘medically treated’ and were further subdivided into rate or rhythm control groups, consistent with a prior SAGE-AF analysis [15]. In brief, rhythm control was defined as the use of an antiarrhythmic drug or prior cardioversion, while rate control included all SAGE-AF participants who did not report treatment with an antiarrhythmic drug or cardioversion during their baseline examination.

### 2.4 Clinical Outcomes

The pre-specified analysis of major adverse cardiovascular endpoints (MACE) included mortality due to cardiovascular cause (vascular death), myocardial infarction (MI), stroke, deep vein thrombosis (DVT), pulmonary embolism (PE), and major bleeding. These events were obtained from participants’ medical records, death certificates, and follow-up assessments. Major bleeding events were adjudicated by a physician committee and classified according to the International Society of Thrombosis and Hemostasis scale [16]. The major bleeds were fatal, occurred in a major organ, or required more than 2 units of transfusion due to hemoglobin loss [17]. A subgroup analysis was performed in both groups to assess the effects of warfarin versus DOACs on cognitive function.

### 2.5 Quantitative Variables and Statistical Methods

Our analytical sample of 887 included those who had completed cognitive function evaluations (described below) at baseline, 1-year, and 2-year follow-ups. Demographic and baseline clinical characteristics of the participants were compared between participants with a previous ablation procedure at the beginning of the study and participants without a history of ablation using analysis of variance for continuous variables and the Chi-square test for categorical variables. All individuals in the CA group underwent the procedure prior to enrollment in the study.

A propensity score for undergoing CA was calculated for each participant in this analysis. Variables that were included in the propensity score were sex, age, BMI, CHA_2_DS_2_-VASc score, AF type (paroxysmal, persistent, or permanent), education level, smoking status, history of hypertension, history of heart failure, history of diabetes, history of stroke, history of renal disease, history of sleep apnea, use of rhythm control drugs, use of rate control drugs, and use of oral anticoagulants. These variables were selected based on clinical relevance and significance in **Table 1**. Mixed-effects logistic regression was employed for longitudinal binary response data to examine the relationship between prior ablation and cognitive impairment. This relationship was adjusted for the propensity score, maintenance of normal sinus rhythm (NSR) as an indication of CA success, and treatment group (rhythm vs rate control). The relationship between the type of anticoagulation treatment and cognitive impairment was adjusted for the propensity score only.

**Table 1.** Participant characteristics according to receipt of catheter ablation procedure for AF prior to enrollment.

| Characteristics | Catheter Ablation Status at Baseline (n) |  |  |
| --- | --- | --- | --- |
|  | Yes (n=193) | No (n=694) | P-value |
| **Socio-demographics** |  |  |  |
| Age, mean, years (SD)* | 73.0 (6.1) | 75.9 (6.8) | **<0.0001** |
| Age, category (%) |  |  |  |
| 65-74 years | 123 (63.7) | 331 (47.7) | **<0.001** |
| 75-84 years | 58 (30.1) | 277 (39.1) |  |
| 85 years or older | 12 (6.2) | 86 (12.4) |  |
| Female sex (%) | 95 (49.2) | 336 (48.4) | 0.84 |
| Race (%) |  |  |  |
| Non-Hispanic White | 174 (90.2) | 601 (86.7) | 0.20 |
| Education (%) |  |  |  |
| College graduate or more | 86 (44.6) | 309 (45.0) | 0.90 |
| AF Type |  |  |  |
| Paroxysmal | 118 (61.1) | 415 (59.8) | **<0.01** |
| Persistent | 60 (31.1) | 156 (22.5) |  |
| Permanent | 4 (2.1) | 41 (5.9) |  |
| Other | 11 (5.7) | 82 (11.8) |  |
| **Baseline (%)** |  |  |  |
| Cognitive Impairment (MOCA ≤23) | 62 (32.1) | 274 (39.5) | 0.06 |
| Frailty |  |  |  |
| Frail | 28 (14.5) | 76 (11.0) | 0.36 |
| Prefrail | 94 (48.7) | 364 (52.5) |  |
| Not Frail | 71 (36.8) | 254 (36.6) |  |
| Able to walk 15 ft | 191 (99.0) | 678 (97.7) | 0.39 |
| Depression | 56 (29.0) | 165 (23.8) | 0.14 |
| Anxiety | 50 (25.9) | 143 (20.6) | 0.11 |
| Social Isolation (MOS<12) | 30 (15.5) | 84 (12.1) | 0.21 |
| Smoking Status |  |  |  |
| Never Smoker | 95 (49.2) | 332 (47.8) | 0.76 |
| Former Smoker | 94 (48.7) | 341 (49.1) |  |
| Current Smoker | 4 (2.1) | 21 (3.0) |  |
| **Bleeding History (%)** |  |  |  |
| Intracranial hemorrhage | 2 (5.6) | 7 (5.7) | 1.00 |
| GI Bleed | 24 (64.9) | 70 (56.9) | 0.39 |
| Major Bleed | 37 (19.2) | 123 (17.7) | 0.64 |
| Heart Failure | 79 (40.9) | 223 (32.1) | **0.02** |
| CAD – MI or Angina | 48 (24.9) | 187 (27.0) | 0.56 |
| Peripheral Vascular Disease | 20 (10.4) | 104 (15.0) | 0.10 |
| Hypertension | 171 (88.6) | 619 (89.2) | 0.81 |
| Type II Diabetes | 52 (26.9) | 179 (25.8) | 0.75 |
| Dyslipidemia | 157 (81.4) | 562 (81.0) | 0.90 |
| Ischemic Stroke | 13 (6.7) | 71 (10.2) | 0.14 |
| Anemia | 61 (31.6) | 207 (29.8) | 0.63 |
| COPD | 52 (26.9) | 163 (23.5) | 0.32 |
| Renal Disease | 37 (19.2) | 192 (27.7) | **0.02** |
| Implantable Cardiac Device | 88 (45.6) | 191 (27.5) | **<0.0001** |
| Sleep Apnea | 65 (33.7) | 181 (26.1) | **0.04** |
| **Blood Levels** |  |  |  |
| Creatinine (mg/dL) | 1.01 (0.29) | 1.08 (0.49) | 0.11 |
| Hemoglobin (mg/dL) | 12.9 (1.8) | 13.2 (1.8) | 0.06 |
| Platelets | 205.6 (63.0) | 210.7 (73.2) | 0.61 |
| **Risk Scores** |  |  |  |
| CHA2DS2-VASc Score (SD)* | 4.24 (1.46) | 4.36 (1.60) | 0.50 |
| HAS-BLED Score (SD)* | 3.15 (1.01) | 3.21 (1.08) | 0.45 |
| **Medications (%)** |  |  |  |
| Total # |  |  |  |
| 1 | 38 (19.7) | 154 (22.2) | 0.41 |
| 2 | 58 (30.1) | 173 (24.9) |  |
| 3 | 41 (21.2) | 173 (24.9) |  |
| 4+ | 56 (29.0) | 194 (28.0) |  |
| Any oral anticoagulant | 173 (89.6) | 585 (84.3) | 0.06 |
| Warfarin | 69 (39.9) | 370 (63.1) | **<0.0001** |
| Antiplatelet | 11 (5.7) | 41 (5.9) | 0.91 |
| Treatment method |  |  |  |
| Rhythm control | 137 (71.0) | 317 (45.7) | **<0.0001** |
| Rate control | 56 (29.0) | 377 (54.3) |  |

Only patients with MOCA scores at all time points are included. \*Age value, CHA2DS2-VASc Score, and HAS-BLED Score reported with no missing data.

We adjusted for clinically relevant and significant Table 1 variables when examining the relationship between prior ablation and clinical outcomes. These components included age, history of CAD, implantable cardiac devices, sleep apnea, use of antiarrhythmics, and use of anticoagulants. Statistical analyses were performed using SAS c9.4 (SAS Institute Inc., Cary, NC).

## RESULTS

A total of 887 SAGE-AF participants were included in this analysis. The total number of participants at baseline are included both at the one- and two-year follow-up period. Participants included in our analysis were on average 75.2 ± 6.7 years old, 48.6% were women, and 87.4% self-identified as non-Hispanic white. Of the total, over half (60.1%) were classified as having paroxysmal atrial fibrillation, and 193 (21.8%) individuals had undergone a CA procedure before enrollment. 336 (37.9%) participants were cognitively impaired at baseline. The mean scores of CHA_2_DS_2_-VASc and HAS-BLED at baseline were 4.33 ± 1.57 and 3.20 ± 1.06, respectively.

Demographic and clinical characteristics by CA history are shown in **Table 1**. Participants who had undergone a CA procedure pre-enrollment were younger and less likely to be taking warfarin *(39.9% vs. 63.1%, p < 0.0001)* or rate control drugs *(80.0% vs 80.6%, p < 0.05)* than were participants in the non-CA group. Of the CA group, 173 (89.6%) were on any OAC at baseline, compared to 585 (84.3%) of the non-CA group (p = 0.06). Those who had undergone a CA were more likely to have persistent AF than participants in the non-CA group (*31.1% vs 22.5%, p < 0.05*). Participants who were treated with CA pre-enrollment were more likely to have also undergone ICD implantation (*45.6% vs 27.5%, p < 0.001*), and were more likely to have comorbid heart failure (*40.9% vs 32.1%, p < 0.05*) and sleep apnea (*33.7% vs 26.1%, p < 0.05*), whereas they were less likely to have chronic renal disease (*19.2% vs 27.7%, p < 0.05*) when compared with those who did not receive CA **(Table 1**).

### 3.1 CA vs. Medical Management and Relation to Cognitive Impairment

There were no significant differences in rates of cognitive impairment at baseline between SAGE-AF participants who had undergone a CA procedure before enrollment vs those who did not (*32.1% vs 39.5%, p = 0.06*). However, participants in the CA group were significantly less likely to be cognitively impaired over the two-year study follow-up, even after inclusion in a model adjusting for propensity to undergo CA (*aOR_1_ 0.70, 95% CI 0.50-0.97*). A second model (Model 2) attenuated the association between CA and cognitive impairment to non-significance by adjusting for AF treatment strategy (rate vs. rhythm control) and maintenance of NSR from baseline to 2-years in addition to propensity score *(aOR_2_ 0.75, 95% CI 0.52-1.08)* **(Table 2)**. **Figure 1** provides a graphics depiction of the MoCA score at baseline, one- and two-year follow-up in both the CA and non-CA groups. The total number of individuals with a MoCA ≤ 23 at baseline, one-, and two-year follow-up are included in **Table 3** and **Figure 2**. At baseline 62 individuals (32.1%) in the CA had cognitive impairment compared to 274 (39.5%) in the non-CA group (*p = 0.06)*. At the two-year end point 58 individuals (30.1%) in the CA group and 311 (44.8%) in the non-CA group had a MoCA ≤ 23 (*p = 0.0002)*. Further longitudinal analysis of mean raw MoCA scores for each group is given in **Supplemental Table 1**. NSR status based on CA at baseline and the 2-year endpoint is provided in **Supplemental Table 2**.

**Figure 1:**
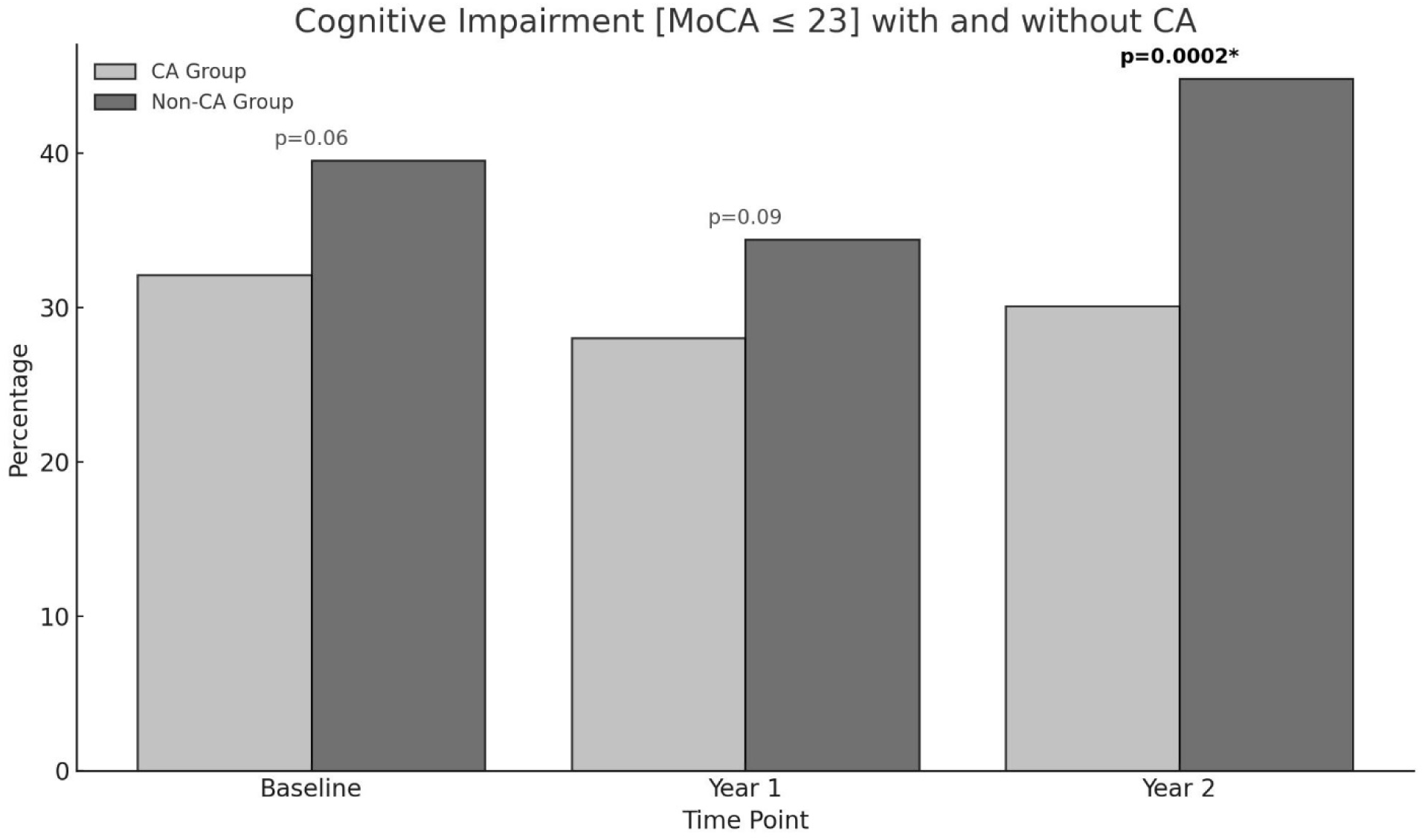
Percentage of individuals in the CA and non-CA groups with cognitive impairment defined by a MoCA ≤ 23 at baseline, one-, and two-year end points. At baseline (p = 0.*06*) and at one-year (*p = 0.09*) no statistically significant difference in cognitive impairment was observed between both groups. At year two a significant proportion of individuals in the non-CA group are cognitively impaired compared to the CA group (*p= 0.0002)*.

**Figure 2:**
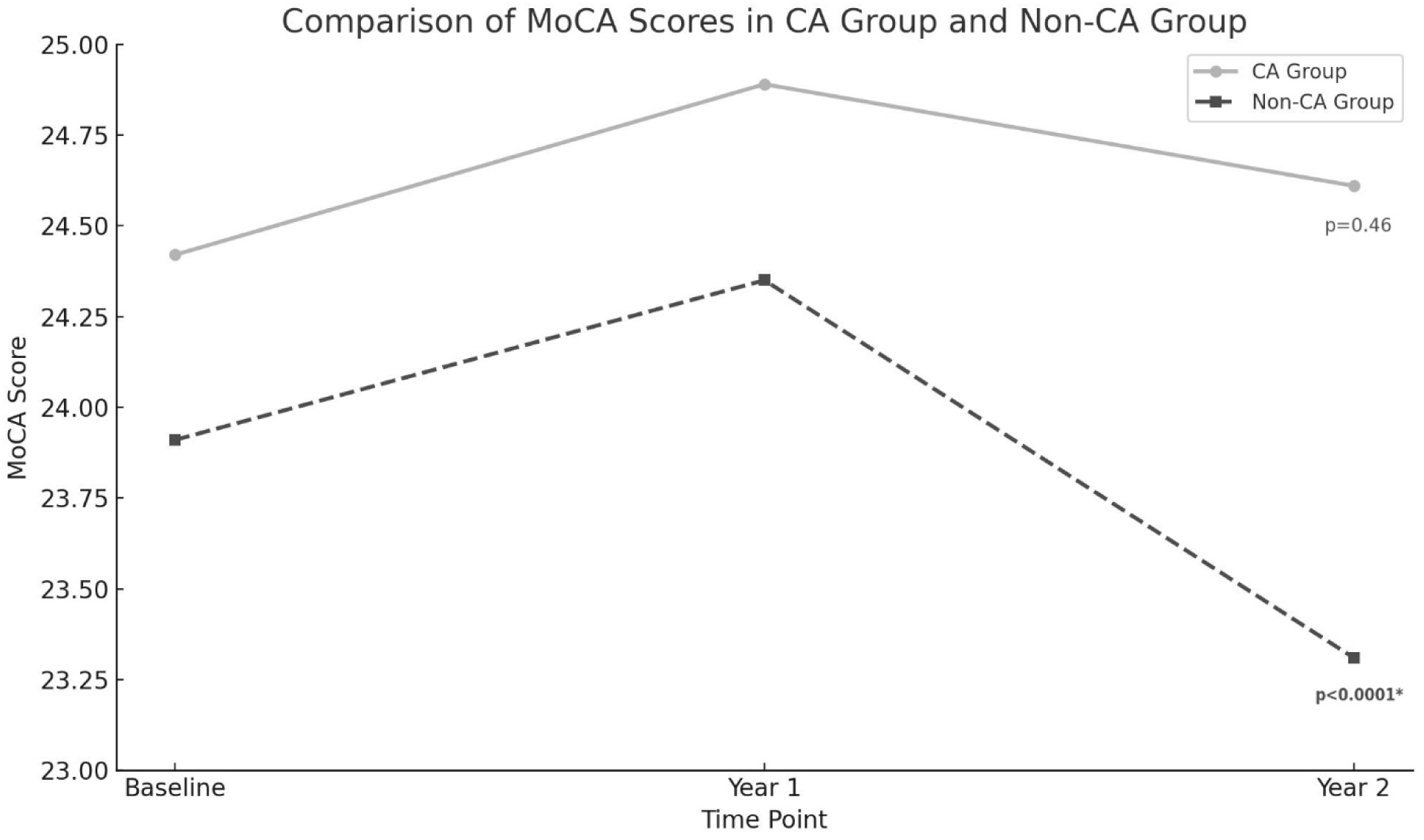
MoCA score at baseline, one, and two-year endpoints in both the CA and non-CA groups. A statistically significant drop in the MoCA score was observed at the two-year end point compared to baseline in the non-CA group (*p <0.0001)*.

**Table 2.**
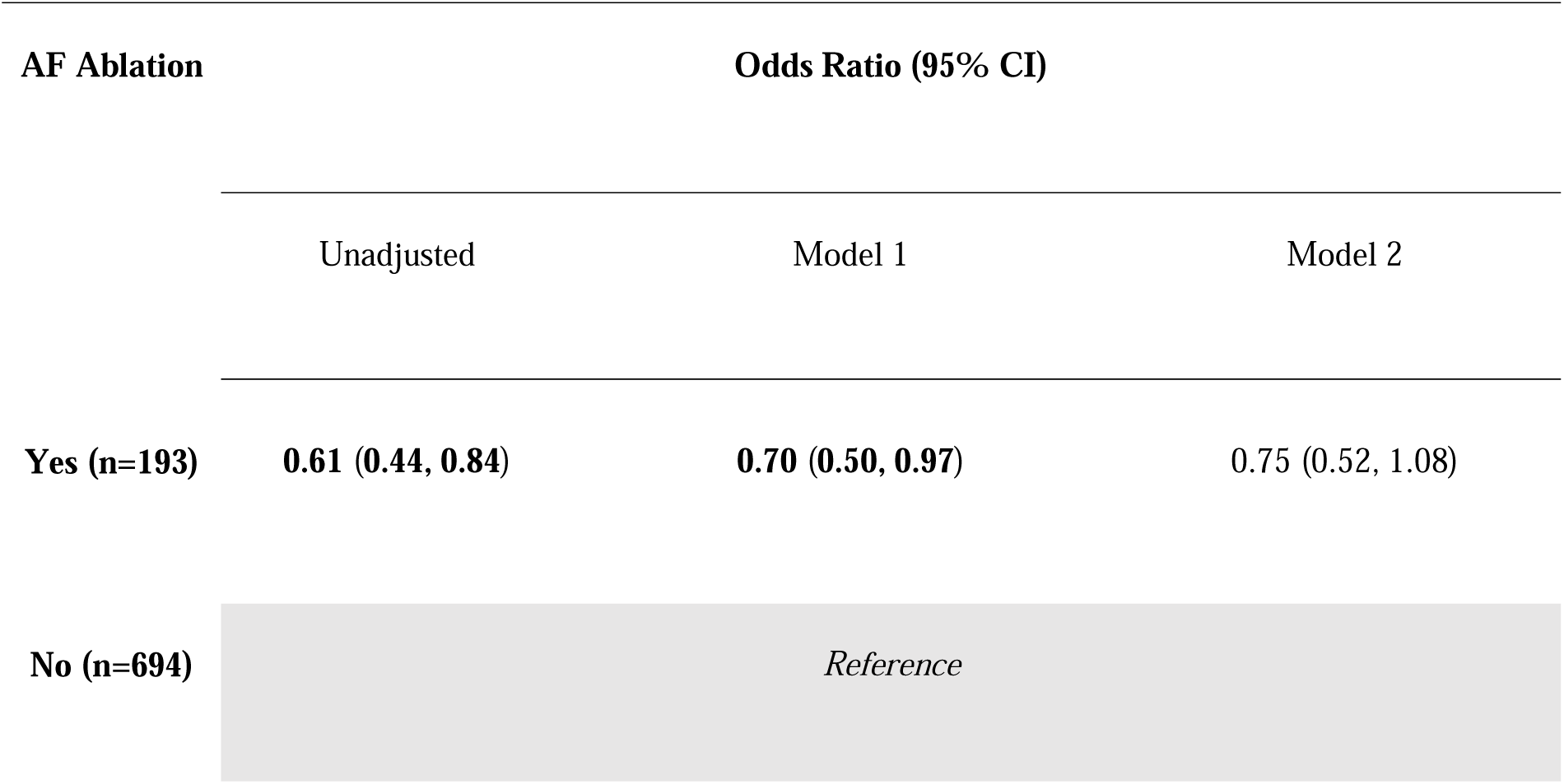
Odds (95% Cis) of cognitive impairment (MoCA<=23) over 2-years by prior catheter ablation for AF status at study enrollment. Model 1 adjusts for propensity score (sex, age, BMI, CHA_2_DS_2_-VASc score, AF type (paroxysmal, persistent, or permanent), education level, smoking status, history of hypertension, history of heart failure, history of diabetes, history of stroke, history of renal disease, history of sleep apnea, use of rhythm control drugs, use of rate control drugs, and use of oral anticoagulants). Model 2 adjusts for Model 1 plus treatment group (rate vs rhythm control) and maintenance of NSR.

**Table 3.** Number of individuals with MoCA ≤ 23 at baseline, one-, and two-year follow-up.

| Cognitive<br>Impairment (MoCA<br>$\leq 23$) | AF Ablation? | | P-Value |
| --- | --- | --- | --- |
|  | Yes (N=193) | No (N=694) |  |
| **Baseline** | 62 (32.1) | 274 (39.5) | 0.06 |
| **Year 1** | 54 (28.0) | 239 (34.4) | 0.09 |
| **Year 2** | 58 (30.1) | 311 (44.8) | **0.0002** |

### 3.2 Major Ischemic/Hemorrhagic Events in CA and Medically Managed AF Patients

After adjusting for confounding variables, we noted no statistically significant association between prior CA and odds of major bleeding (*aOR 0.53, 95% CI 0.24-1.16*) or the pre-specified composite outcome of all-cause MACE (ischemic stroke, vascular death, MI, and major bleeding) (*aOR 0.65, 95% CI 0.35-1.20*) over 2 years follow-up (**Supplemental Table 3**).

### 3.3 Subgroup Analysis of Warfarin vs. DOACs on Cognitive Impairment

We did not observe any significant difference in either the CA or the no CA groups between the use of warfarin vs DOACs on cognitive function, even after adjusting for propensity to receive CA (**Table 4**). Notably, both the CA and non-CA groups had high rates of oral anticoagulant use, with nearly 90% of the CA group and 84% of the non-CA group being prescribed an oral anticoagulant throughout the study period.

**Table 4.** Odds of cognitive impairment over 2-years using Warfarin vs. all other AC. Model 1 adjusts for propensity score (sex, age, BMI, CHA_2_DS_2_-VASc score, AF type (paroxysmal, persistent, or permanent), education level, smoking status, history of hypertension, history of heart failure, history of diabetes, history of stroke, history of renal disease, history of sleep apnea, use of rhythm control drugs, use of rate control drugs, and use of oral anticoagulants). Model 2 adjusts for Model 1 plus treatment group (rate vs rhythm control) and maintenance of NSR.

| Ablation<br>Group | Odds Ratio (95% CI) |  |  |
| --- | --- | --- | --- |
|  | Unadjusted | Model 1 | Model 2 |
| **CA**<br>**(n=193)** | 0.80 (0.43, 1.48) | 0.78 (0.41, 1.46) | 0.77 (0.41, 1.43) |
| **No CA**<br>**(n=694)** | 1.07 (0.76, 1.50) | 1.02 (0.72, 1.43) | 0.97 (0.69, 1.36) |

## DISCUSSION

### 4.1 Atrial Fibrillation and Cognitive Decline

In this retrospective analysis of an observational study that performed a longitudinal assessment of cognitive function, we found that participants who had undergone CA for AF at baseline were less likely to develop cognitive impairment over a 2-year follow-up compared to those who underwent medical rhythm or rate control, even after adjusting for propensity to undergo CA. These results are consistent with prior studies associating the persistence of AF with cognitive impairment, including an analysis that examined data from both the ONTARGET and TRANSCEND trials in 2012. In this study, the authors showed that AF was associated with an increased risk of cognitive decline *(HR 1.14, CI 1.03-1.26*), and incident dementia (*HR 1.30, CI 1.14-1.49*) [18]. Our study is one of the few to demonstrate a link between CA and cognitive function among older AF patients using serial objective measures of cognitive health.

AF is hypothesized to cause cognitive impairment primarily through cerebrovascular accident and cerebral microbleeds among other causes. Possible mechanisms include decreased cerebral blood flow, impaired autoregulation, endothelial dysfunction, and chronic inflammation [19]. Disruptions in the former occur in AF and have been linked with a decline in cognitive function in patients with AF with or without a history of stroke [20, 21]. In 37,025 prospectively analyzed patients without AF for 5 years, 10,161 (27%) developed AF and 1,535 (4.1%) developed dementia. AF has been independently associated with developing dementia with an increased risk of mortality [22–25].

It has been hypothesized that effective oral anticoagulation will therefore improve cognitive outcomes [26]. Rhythm control through pharmacologic intervention or ablation has also been theorized to have benefits in cognition in AF through the prevention of rapid ventricular response [27]. As mentioned, the non-CA group was more cognitively impaired at the 2-year endpoint compared to the CA group. These results are consistent with the AFFIRM study [28]. Furthermore, there was no significant increase in both obvious ischemic/hemorrhagic events in patients who underwent CA. The major hypothesis for the observed cognitive benefit is due to a favorable hemodynamic profile in the cerebral microvasculature. Prior studies from the SAGE-AF cohort demonstrated a 9% comorbidity between frailty and cognitive impairment [29]. We did observe a non-significant decrease in cognitive impairment in the CA group when controlling for NSR in addition to propensity matching (**Table 2**).

One study of cognitive outcomes utilizing MoCA score in AF patients who underwent ablation compared to a propensity-matched group who did not undergo ablation found that CA improved performance at 1 year. Similarly, in our cohort, a slightly higher MoCA score at the 2-year endpoint in the CA group (*24.42±3.67 vs. 24.61±3.71*). Another study utilized near-infrared spectroscopy to identify regional cerebral blood flow and cerebral activity in patients with persistent AF compared to post-ablation AF patients who maintained sinus rhythm found CA improved frontal and temporal brain activities in some patients and was associated with an improvement in depression and cognitive function [30, 31]. Our results, in conjunction with previous studies, can help guide future research in identifying definitive biological mechanisms for the neurocognitive benefits of ablation.

### 4.2 Warfarin and OACs on Cognitive Function

The subgroup analysis revealed no difference in cognitive outcomes between those on warfarin and all other OACs. This result was observed in both AF patients who underwent CA and those who were only medically managed. Previous studies have shown benefits in the utilization of DOACs over warfarin, thought to be due to warfarin’s propensity to cause cerebral microbleeds (CMBs) and silent infarction. Studies comparing warfarin and DOACs reported that patients on DOACs for AF had a lower incidence rate of dementia when compared to warfarin. In our study, both groups were treated highly with anticoagulation due to their advanced age putting them at higher CHA_2_DS_2_-VASc scores. There was a higher proportion of patients receiving warfarin in the no-CA group (*63.1% vs 39.9%*). This could be interpreted to drive the elevated bleeding rates (*7.5% vs 4.2%*) and MACE (*10.4% vs 6.2%*) observed in the no-CA vs CA groups, which was underpowered for significance. Our subgroup analysis argues against warfarin as a cause of elevated MACE, CMBs, or silent infarction as there was no difference in cognitive outcomes. Further prospective head-to-head comparative studies between specific anticoagulants are needed.

### 4.3 Strengths and Limitations

The main strengths of our research are that the data were collected prospectively over 2 years. Furthermore, a validated standardized instrument (MoCA score) was used to examine cognitive function, enhancing reproducibility and sensitivity to subtle changes in cognitive function. However, our study also has several limitations. Our cohort included a smaller proportion of patients who underwent CA compared to those who were medically managed. Furthermore, we did not have a high number of participants who underwent medical rhythm control. We did not distinguish between the specific types of CA procedures or the number of recurrent CA procedures. Participants in our study were primarily non-Hispanic white and well-educated, which may impair the generalizability of our findings to other racial or ethnic groups or less well-educated AF patients. We were unable to compare interactions between CA, warfarin, and cognitive health in comparison to each DOAC as there were insufficient numbers of DOAC-treated patients in each treatment group. Observational studies detect associations and cannot establish causal relations between CA and cognitive health. However, we supported our observations with biologically plausible mechanisms and conducted rigorous propensity score adjustment.

## CONCLUSIONS

Catheter ablation is widely performed to improve symptoms and reduce AF burden. Our findings suggest that adults with AF who undergo catheter ablation are also less likely to become cognitively impaired than those who receive medical treatment alone over a two-year follow-up period. This is not explained by differences in rates or types of oral anticoagulants used between groups or differences in the characteristics of participants at baseline. Further contemporary studies including randomized controlled trials are needed to validate our findings and examine whether ablation technique (e.g., radiofrequency, cryothermal, pulse field), duration of the procedure or post-procedure anticoagulation use, or cerebral hemodynamics further influence cognition after CA among patients with AF.

## Data Availability

All data produced in the present study are available upon reasonable request to the authors

