## Supplemental Tables for "Is Catheter Ablation Associated with Preservation of Cognitive Function? An Analysis From the SAGE-AF Observational Cohort Study"

### Supplemental Table 1. Cognitive function by prior CA for AF status among SAGE-AF participants at Baseline, 1-year, and 2-year follow-up examinations.

|  | **Ablation (N=193)** | | **No Ablation (N=694)** | |
| --- | --- | --- | --- | --- |
|  | MoCA Score (SE) | P-Value | MoCA Score (SE) | P-Value |
| Baseline | 24.42 (3.67) | 0.46 | 23.91 (3.72) | **<0.0001** |
| Year 1 | 24.89 (3.27) |  | 24.35 (3.87) |  |
| Year 2 | 24.61 (3.71) |  | 23.31 (4.16) |  |

### Supplemental Table 2. Normal sinus rhythm by prior CA status at baseline and 2-year follow-up examination.

|  | AF Ablation | |  |
| --- | --- | --- | --- |
|  | Yes (n=162) | No (n=631) | P-Value |
| Normal sinus rhythm |  |  |  |
| Maintenance of NSR (both Baseline and Year 2) | 71 (43.8) | 158 (25.0) | **<0.0001** |
| Baseline only | 14 (8.6) | 67 (10.6) |  |
| Year 2 only | 21 (13.0) | 110 (17.4) |  |
| Never NSR | 56 (34.6) | 296 (46.9) |  |

### Supplemental Table 3. Odds (95% CIs) of developing a major bleeding event or any major adverse cardiovascular endpoint (stroke, vascular event, MI, and major bleeding) by CA status at baseline.

| **Variables** | **Major Bleed** | **Composite Outcome** |
| --- | --- | --- |
| Outcome among CA group | 8 (4.2%) | 14 (6.2%) |
| Outcome among the non-ablated group | 56 (7.5%) | 78 (10.4%) |
| Crude OR (95% CI) | 0.49 (0.23-1.05) | 0.56 (0.30-1.06) |
| Adjusted* OR (95% CI) | 0.53 (0.24-1.16) | 0.58 (0.30-1.11) |
| *Adjusted for age, AF type, CAD, ICD, sleep apnea, anticoagulation treatment, and rate-control treatment.  The composite outcome includes ischemic stroke, vascular mortality, MI, and major bleeding. | | |
